# Treatment of left ventricular dysfunction in dilated cardiomyopathy with a myeloperoxidase inhibitor

**DOI:** 10.1101/2022.11.02.22281785

**Authors:** Simon Geissen, Simon Braumann, Joana Adler, Felix Sebastian Nettersheim, Dennis Mehrkens, Alexander Hof, Henning Guthoff, Philipp von Stein, Sven Witkowski, Norbert Gerdes, Lea Isermann, Aleksandra Trifunovic, Alexander C. Bunck, Martin Mollenhauer, Holger Winkels, Matti Adam, Anna Klinke, Martin Hellmich, Malte Kelm, Volker Rudolph, Stephan Rosenkranz, Stephan Baldus

## Abstract

Dilated cardiomyopathy (DCM), an incurable disease of the cardiomyocyte terminating in systolic heart failure (HFrEF), is prevalent, causes hospitalization and is associated with increased mortality. Despite evidence of immune activation in DCM, anti-inflammatory interventions so far did not prove to alter the course of this disease. Here we show that myeloperoxidase (MPO), the principal heme peroxidase expressed by polymorphonuclear neutrophils (PMN) and monocytes, critically contributes to HFrEF in DCM.

Muscle LIM protein (MLP) deficient mice, which spontaneously develop DCM, display increased circulating PMN counts and augmented levels of vessel-immobilized MPO. Genetic ablation and pharmacological inhibition of MPO resulted in enhanced nitric oxide (NO) bioavailability of systemic conductance and resistance vessels, and subsequently restoration of systolic left ventricular (LV) function, whereas infusion of MPO worsened systolic LV function. When patients diagnosed for DCM were treated with an orally available MPO inhibitor, systolic LV function increased, natriuretic peptides declined, and functional status improved.

Impairment of endothelial NO bioavailability by release of leukocyte-derived MPO evolves as a disease-aggravating mechanism in DCM. MPO inhibition profoundly improved ventricular function by lowering systemic vascular resistance and thus holds promise as a novel and complementary treatment strategy for patients with DCM.

## Introduction

Heart failure with reduced ejection fraction (HFrEF) is the leading cause of cardiovascular hospitalization in Western countries, accounts for an annual mortality of nearly 10% and - despite recent medical progress – remains uncurable.^1^ Pharmacological interventions, which lower mortality in HFrEF, proved beneficial by unloading the ventricle rather than by augmenting inotropy: Beta receptor-blocking agents blunt sympathetic overstimulation, inhibitors of the renin angiotensin system (RAS) and of neprilysin decay reduce systemic vascular resistance and mineralocorticoid receptor antagonists and SGLT2 inhibitors lessen myocardial fibrosis and fluid retention, respectively.

Pathophysiologically, HFrEF is accompanied by inflammatory processes: Leukocyte activation, increase in cytokines and reactive oxygen species (ROS) and impaired nitric oxide (NO) bioavailability have been firmly linked to reduced left ventricular ejection fraction (LVEF).^2^ However, whether inflammation is cause or consequence of heart failure so far remains elusive, whereas anti-inflammatory treatment strategies have largely failed to alter the course of this disease.^3^ Myeloperoxidase (MPO), a heme protein abundantly expressed by and released from neutrophils (PMN) and monocytes, functions as a constituent of the innate immune defense but also demonstrated to affect ventricular remodeling following ischemic injury: Depletion or pharmacological inhibition of MPO in animal models of myocardial infarction improved left ventricular function and ameliorated ventricular dilatation, which was linked to decreased recruitment of PMN and blunted matrix metalloproteinase activation.^4,5^ Similarly, MPO deficient mice were protected against angiotensin II (Ang II)-mediated atrial fibrosis and fibrillation.^6^

Myeloperoxidase has also been shown to affect vascular tone: MPO binds to endothelial cells and oxidizes NO – both directly and via generation of small radical intermediates - in the subendothelial space.^7,8^ Oxidation of the NO synthase (NOS) product L-arginine and inactivation of dimethylaminohydrolase (DDAH), which increases the bioavailability of endogenous NOS inhibitors, complement the ability of MPO to increase vascular tone by reducing endothelial NO bioavailability.^8,9^ It was proposed that MPO inhibition enhances cyclic guanosine monophosphate (cGMP) levels in diseased vessels by reducing oxidation of soluble guanylate cyclase (sGC).^10^ Thus, MPO emerges as an enzyme capable of increasing vascular tone by decreasing NO bioavailability.

Dilated cardiomyopathy (DCM) is a principal cause of HFrEF and in relevant part based on genetic defects in the contractile apparatus of the cardiomyocyte. Muscle LIM protein (MLP)-deficiency (*Mlp*^*-/-*^) serves as a well-characterized murine model of DCM with development of left ventricular dilation and reduced LVEF irrespective of an inflammatory insult and devoid of obvious stimuli of neutrophil activation. We thus used this mouse model to investigate whether MPO contributes to the initiation and progression of systolic heart failure.^11,12^

## Materials and methods

A detailed methods section can be found in the supplementary material.

### Animal model

The generation of *Mlp*^*-/-*^ mice has been described previously.^12^ Here, MPO deficient animals^13^ were backcrossed to the FVB/N background and consecutively, the *Mlp*^*-/-*^ and *Mpo*^*-/-*^ lines were cross-bred to generate animals that are deficient for both proteins. All mice experiments were conducted in male mice and controlled in order to exclude potential litter effects. All animal experiments were conducted under permission of the Landesamt für Natur-, Umwelt-und Verbraucherschutz Nordrhein-Westfalen, Recklinghausen, license numbers 2015.A459 and 2017.A464.

### Treatment of DCM patients with AZD4831

Patients with documented DCM and HFrEF, who were on stable guideline-directed HF therapy for at least 12 weeks, were treated with AZD4831 (5 mg once daily) for 12 weeks. In order to qualify for study inclusion, all patients underwent right heart catheterization. The included patients displayed postcapillary pulmonary hypertension (mPAP 28 ± 4.1 mmHg, PVR 2.5 ± 0.49 WE). All patients underwent clinical assessment including physical examination, New York Heart Association (NYHA) functional class grading, assessment of the six-minute walking distance (6MWD) and NTproBNP serum levels (Elecsys proBNP II Test, Roche Diagnostics GmbH, Mannheim, Germany) at baseline, and at regular follow-up visits up to week 12. Furthermore, all patients underwent repeat cMRI at baseline and at week 12. The main objective was to assess the preliminary efficacy of 12 weeks of AZD4831 administration on clinical measures (NTproBNP levels, 6MWD) and LVEF in patients with HFrEF (DCM). The main safety objective was to evaluate the safety and tolerability of multiple doses of AZD4831 in DCM patients. The study was approved by the local Ethics Committee of the Medical Faculty of the University of Cologne (No. 19-1612-AMG-ff). The study is registered as EudraCT-No. 2020-002788-80.

### Statistical analysis

For comparison of apparently normal-distributed measures between two groups, Student’s t-test was calculated. For comparison of three or more groups, one-way ANOVA followed by Tukey’s post-hoc comparisons were used. Since the distribution of NTproBNP levels is usually positively skewed, logarithms were taken first and changes over time were analyzed with the paired t-test (this corresponds to the ratio-paired t-test on original scale). All calculations and plotting were done using GraphPad Prism, Version 9 (GraphPad Software, San Diego, CA, USA) unless otherwise indicated.

### Data availability

All data supporting the findings of this study are available from the corresponding author upon request.

## Results

To assess the innate immune response in the context of DCM, PMN counts and vascular MPO deposition were analyzed. At 14 weeks, *Mlp*^*-/-*^ mice had augmented circulating PMN counts and increased levels of vessel-immobilized MPO upon liberation by heparin perfusion (Figure 1a-b). Since PMN recruitment and activation in DCM was observed in the absence of cardiomyocyte necrosis (Supplemental figure 1a), we tested if activation of the neurohumoral RAS, a paramount event in systolic heart failure,^14^ is the potential cause for enhanced MPO levels. Indeed, we found increased circulating levels of Ang II in *Mlp*^*-/-*^ mice (Figure 1c). Furthermore, whole blood samples challenged with physiological levels of Ang II revealed MPO release from PMN comparable to lipopolysaccharide (LPS; Figure 1d).

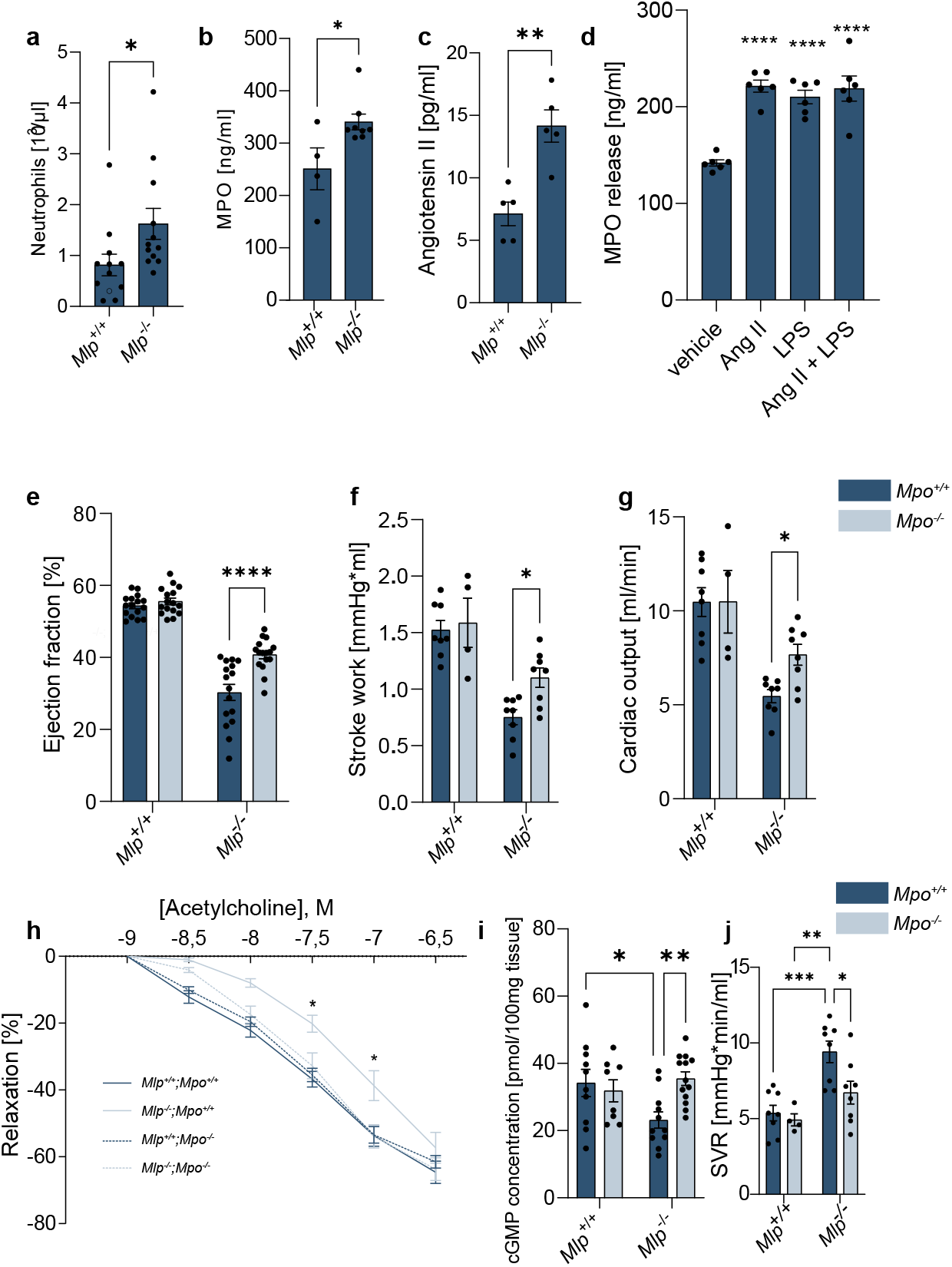

In order to characterize the potential role of MPO as a modifier of the cardiac phenotype in DCM, *Mlp*^*-/-*^ mice were crossbred with MPO deficient (*Mpo*^*-/-*^) animals (*Mpo*^*-/-*^/*Mlp*^*-/-*^). At 14 weeks of age, *Mlp*^*-/-*^ mice had developed a significant decline in systolic LV function. In contrast, *Mlp*^*-/-*^*/ Mpo*^*-/-*^ animals revealed significantly improved LVEF as compared to MPO competent *Mlp*^*-/-*^ mice. Pressure-volume (PV) loop assessment of the left ventricle revealed increased stroke work and cardiac output (Figure 1d-f).

Given the profound oxidative effects of MPO, we next investigated left ventricular remodeling in *Mlp*^*-/-*^ and *Mlp*^*-/-*^*/Mpo*^*-/-*^ mice: Using electron microscopy, we found no difference in sarcomeric ultrastructure in cardiomyocytes of the two groups (Supplemental figure 1b). Also, total collagen content, an indicator for fibrotic remodeling, was not altered in the absence of MPO (Supplemental figure 1c). Moreover, neither mitochondrial stress response nor complexes of oxidative phosphorylation were affected by MPO, indicating unaltered mitochondrial function (Supplemental figure 2-3). Also, MPO had no impact on cMyBP-C phosphorylation in *Mlp*^*-/-*^ mice (Supplemental figure 2-3). Finally, ex vivo infusion of MPO did not affect inotropy in Langendorff perfusion of explanted hearts (Supplemental figure 1d-f). All in all, we did not find any evidence for an adverse effect of MPO on cardiomyocyte contractility and myofibroblast activation, respectively.

In contrast, we observed a strong MPO-driven vascular phenotype: Explanted aortic rings of *Mlp*^*-/-*^/*Mpo*^*-/-*^ in comparison to *Mlp*^*-/-*^/*Mpo*^*+/+*^ animals displayed improved relaxation in response to acetylcholine (Figure 1g), illustrating increased NO bioavailability in the absence of MPO. Also, cGMP levels were increased in MPO naïve aortic tissue (Figure 1h). Consequently, peripheral vascular resistance in the absence of MPO was markedly attenuated (Figure 1i).

To further determine a mechanistic role of MPO in the progression of DCM, recombinant MPO (daily dose: 12.5 pg/g/min body weight) was intravenously infused into *Mlp*^*-/-*^*/Mpo*^*-/-*^ mice for seven days using minipumps. MPO infusion provoked a significant decline in LVEF (Figure 2a). Intravenous infusion of MPO via the tail vein yielded a similar yet instantaneous reduction of LVEF, further supporting the notion of a vascular-humoral rather than a myocardial-structural effect of MPO in this disease (Figure 2b). A single in-vivo injection of MPO in mice rapidly increased systemic vascular resistance (SVR) and diminished LVEF (Figure 2b and c). To test whether MPO inhibition has the potential to improve LVEF in-vivo, we fed *Mlp*^*-/-*^ mice an orally active MPO inhibitor, AZM198. Treatment of mice with this compound for seven days resulted in a significant improvement of LVEF (Figure 2d).

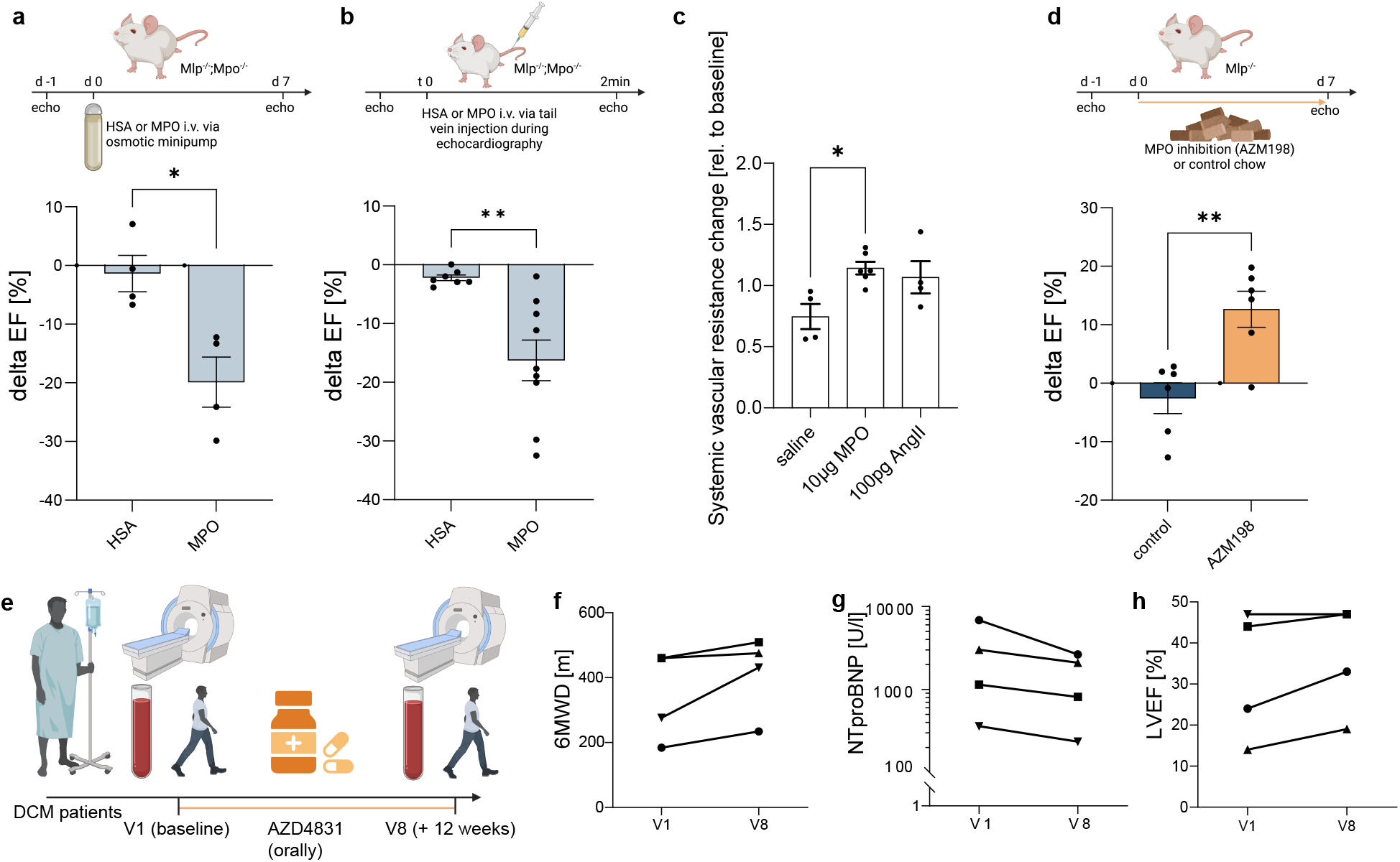

Finally, we tested whether these observations can be translated into human pathology. We therefore conducted an investigator-initiated, open-label phase 2a pilot study approved by the German Federal Institute for Drugs and Medical Devices (BfArm) using the oral MPO-inhibitor AZD4831 in this disease: Five patients diagnosed with DCM (four male, mean age 64.6 ± 6.1 years, mean ejection fraction 33.0 ± 12.4%, full patient characteristics in Supplemental table 1) and under optimal heart failure therapy were treated with this compound. HF-related medication remained unchanged over the course of the study (Supplemental table 2). One patient discontinued the intake at day 5 due to development of a rash, the remainder received the compound for 12 weeks without adverse reactions. At week 12, the 4 patients revealed an increase in 6-minute-walking distance (+ 67.3 ±26.3 m), a significant decline in NTproBNP serum levels (−1390.3 ±821.2 U/l) and an increase in MRI-based LVEF (+ 4.3 ±1.6 %), Figure 2f-h). Extensive serum proteomics analysis confirmed significant reduction of NTproBNP levels from baseline to end of treatment. At the same time 169 other proteins – among those Interleukin (IL)-10 and IL-13 - were found to be significantly regulated in human serum after 12 weeks of treatment with AZD4831 (Supplemental figure 7).

## Discussion

Here we show that leukocytes promote systolic heart failure with MPO exerting its detrimental effects by increasing peripheral vascular resistance. Genetic ablation and pharmacological inhibition of MPO improves systolic LV function in a murine model of heart failure and augments functional status in humans with HFrEF.

Mismatch between oxygen supply and demand as in HFrEF triggers a broad spectrum of pathways to counteract central hypoperfusion: Endogenous catecholamines and the autonomous nervous system increase inotropy of the heart and augment peripheral vascular tone - as do vasoactive peptides such as Ang II, endothelin and vasopressin. ROS, e.g. superoxide, impair endothelial NO bioavailability and thereby further increase peripheral resistance.^2^ Ultimately this fuels the increase in SVR, a hallmark in HFrEF and propagator of disease.^15^

We were able to demonstrate, that leukocyte-derived MPO evolves as a promising pharmacological target in this disease – irrespective of its well established adverse effects on myocardial homogeneity and remodeling in animal models of myocardial ischemia.^16–18^ MPO rapidly binds to and is transcytosed by endothelial cells;^19^ indeed, we not only found significant leukocytosis – a finding previously seen in HFrEF patients^20^ - but also increased levels of vessel-immobilized MPO and Ang II in our animal model. We show that Ang II, a principal and long accepted effector in HFrEF, activates PMN and induces release of MPO in a cytokine-like fashion (Figure 1c-d). Since MPO is capable of lowering endothelial NO bioavailability by multiple pathways, this enzyme evolves as a critical modulator of vascular tone in-vivo.^8,21,22^

So far, anti-inflammatory treatment strategies have yielded - if any - modest effects on the course of systolic heart failure and DCM: antibodies directed against TNF-α did not change the course of this disease.^23,24^ Interference with IL-1 signaling did have a modest effect on cardiovascular outcomes in heart failure,^25,26^ however these effects faded in patients with HFrEF of non-ischemic origin.^27^ Finally, neither methotrexate nor colchicine had a substantial impact on LV function and symptoms in HFrEF patients.^3^

Our data reveal that PMN increase vascular resistance by release and endothelial accumulation of NO-oxidizing MPO. Thus, PMN exert additional biologic functions as circulating modulators of vascular tone. From an evolutionary perspective, this may be advantageous to respond to acute bacteremia and counteract exuberant NO dependent peripheral vasodilation. However, in the presence of left ventricular dysfunction, chronically increased afterload augmented by continuous vascular deposition of MPO appears to be detrimental. This maladaptive innate immune response potentially aggravates systolic heart failure, and its inhibition warrants larger scale investigation as a complementary, anti-inflammatory and non-cardiomyocyte centered treatment strategy in DCM and presumably other etiologies of systolic heart failure.

## Acknowledgment

We thank Malin Aurell, Ralph Knöll and Erik Michaëlsson and Klaus Ley for their expert advice and support.

