## Supplemental_material for "Treatment of left ventricular dysfunction in dilated cardiomyopathy with a myeloperoxidase inhibitor"

#### Additional materials and methods

##### Angiotensin II treatment

12week old C57Bl6/J mice of indicated genotypes were treated with 1000ng/kg/min Angiotensin II via implantation of osmotic minipumps (Alzet, Cupertino, CA, USA, Model 1002). In brief, mice were anaesthetised by isoflurane and after formation of a subcutaneous cavity, a pre-filled pump was implanted. The pump was renewed after two weeks of treatment. 21 days after implantation, echocardiography was performed as stated above to assess left ventricular systolic function.

##### Isolated mouse heart perfusion model

To investigate the role of MPO on the heart function and contractility *ex vivo* we used a Langendorff model[1] and modified it as follows: hearts of $Mlp^{-/-}/Mpo^{-/-}$ mice were explanted and mounted into a Langendorff-apparatus (Hugo Sachs, March-Hugstetten, Germany). Once mounted hearts were constantly perfused with Krebs Henseleit Buffer (KHB) and paced to a heart rate of 600 beats/min, while coronary flow was registered using an ultrasound flow probe (Hugo Sachs). Hearts underwent a stabilization period for 20 minutes, followed by 20 seconds of global zero flow ischemia to test coronary reserve and 5 minutes of recovery. Before application of MPO isolated from human leukocytes (50nM) or HSA (50nM) in KHB buffer via a side arm using a syringe driver, baseline parameters including left ventricular developed pressure (LVDP) and coronary flow were measured. The application was stopped after 5 minutes, followed by regular perfusion and monitoring for 30 minutes. In the following, this type of application and monitoring was repeated twice for functional controls. First using 100nM verapamil (Hexal AG, Holzkirchen, Germany) as negative inotropic agent, followed by 12pM isoproterenol (Merck, Taufkirchen, Germany) as positive inotropic compound to confirm that hearts' responsiveness.

#### Echocardiography

At 15 weeks of age, animals were anesthetized with 5% isoflurane and kept under anesthesia with 2% isoflurane. Under continuous monitoring of vital signs, parasternal long-axis and short-axis views were recorded in B-Mode, M-Mode and ECG-triggered kilohertz visualization (EKV) using a Vevo 3100 device and an MX550S transducer (both VisualSonics, Fujifilm, Tokyo, Japan). B-Mode images of the parasternal long-axis were analyzed using Vevo Lab v5 (VisualSonics) by planimetry of end systolic and end diastolic state of the left ventricle. Examiners were blinded against genotypes and treatments during acquisition and analysis. For the indicated experiments, injections were performed via tail vein catheterization.

#### Pressure-Volume-Loop Analysis

For invasive hemodynamic characterization, animals were anesthetized with 5% isoflurane and kept under 1.5-2% of isoflurane for the remainder of the experiment. Under ventilation, both the right carotid artery and the left caval vein were punctured and pressure catheters were inserted to monitor arterial pressure and central venous pressure. Subsequently, a pressure-volume (model SPR-839, Millar, Houston, Texas, USA) catheter was inserted into the left ventricular cavity. After hemodynamics stabilized, baseline measurements were recorded for 20 minutes with specialized acquisition hardware and software (PowerLab and LabChart 7 by ADInstruments, Sydney, Australia). For the indicated experiments, injections were performed via teil vein catheterization and hemodynamic reaction was monitored for the consecutive 15 minutes. All animals which underwent this procedure were euthanized in deep narcosis. Bolus injections of 15% saline and final blood withdrawal were used for saline and cuvette calibration of the volume catheter as described before.

#### MPO injection

MPO isolated from human leukocytes (Planta, Vienna, Austria) or human serum albumin (HSA, Sigma-Aldrich, St. Loius, MO, USA) were injected into the tail vein in concentrations described previously.[2] In short, during the indicated experiment, the tail vein was punctured with a 30G needle connected to a microtube. 50µl of a PBS solution containing 0.3µg/µl of either MPO or HSA was injected by hand as a bolus.

#### Osmotic minipumps

For chronic MPO infusion, MPO isolated from human leukocytes was filled in osmotic minipumps (Model 1007D, Alzet, Cupertino, California, USA). The pumps were equipped with a catheter tube that was prefilled with the same MPO solution. Under isoflurane anesthesia, the left jugular vein was punctured and the catheter inserted and secured with multiple ligatures. After deposition of the pump in the left flank of the mouse, animals were monitored for 7 days. Upon organ harvest, the correct positioning of the catheter was verified. The MPO dosage was 12.5pg/g body weight per min as described before;[3] human serum albumin in the same dosage was used as a control.

#### Inhibitor treatment

$Mlp^{-/-}$ mice and their wildtype littermates were fed with standard chow that had been supplemented (1,146g/kg) with of AZM198, an experimental inhibitor of MPO for murine usage that was provided by AstraZeneca, Stockholm, Sweden. Identical chow diet without AZM198 was fed to littermates. Echocardiography was performed before and after 7 days of treatment.

#### Organ harvest

Under deep anesthesia and analgesia, the rib cage was opened and the animals were euthanized by final blood withdrawal from the left ventricular cavity and consecutive perfusion with 0.9% saline supplemented with heparin (50IU/ml). Consecutively, hearts and aortas were carefully excised and further processed as indicated respectively.

#### Organ bath

Aortic segments of euthanasized mice were carefully dissected and immediately kept in Krebs-Henseleit buffer on ice. We cut 2-mm segments and mounted them onto steel wires connected to force transducers of a standard organ bath filled with 25 ml Krebs-Henseleit buffer at 37°C and gassed with carbogen. Each ring was equilibrated for 30 minutes and continuously stretched up to a tension of 0.7 g. KCl (80 mM) was added twice to each bath with subsequent washing. Endothelium-dependent relaxation were determined in response to increasing doses of acetylcholine.

#### Stimulation of whole blood samples

Blood freshly collected in EDTA coated microtubes was diluted with three volumes of Roswell Park Memorial Institute medium 1640 (RPMI) and stimulated with 1nM of angiotensin II and/or 1ng/ml E. coli lipopolysaccharide under 37°C and 5% carbon dioxide. After indicated stimulation time, the sample was centrifuged at 1500 x g für 10 minutes and the upper aqueous phase was collected for further analysis.

#### ELISA procedure

For measurement of tissue and plasma levels of either MPO, elastase or cGMP, preconfigured kits were used. In short, MPO quantification was performed by a sandwich ELISA (R & D Systems, Minneapolis, MN, USA, Cat.-No. DY3667 for murine MPO and R & D Systems, Minneapolis, MN, USA, Cat.-No. DMYE00B for human MPO). For elastase quantification Mouse Neutrophil Elastase ELISA Kit (Abcam, Cat.-No. 252356) was used. For cGMP quantification form aortic lysates, tissue was processed as described before[4] and a competitive ELISA was performed in accordance with the manufacturer's protocol (Sigma-Aldrich, Cat.-No. CG201).

#### Isolation of cardiac mitochondria

Freshly collected hearts were immediately transferred into 10 ml of prechilled mito-isolation buffer (MIB) [100 mM sucrose, 50 mM KCl, 1 mM EDTA, 20 mM N-tris(hydroxymethyl)methyl-2-aminoethanesulfonic acid, and 0.2% bovine serum albumin (BSA) free from fatty acids (pH adjusted to 7.2)] supplemented with 1 mg of subtilisin (Sigma-Aldrich) per mg of tissue. Approximately 20 long strokes of a Potter S (Sartorius) homogenizer at 1000 rpm were required for homogenization. After centrifugation (800g, 5 min, 4°C), the mitochondria-containing supernatant was transferred into a fresh tube. Pelleted mitochondria (8500g, 5 min, 4°C) were resuspended in 30 ml of MIB and subjected to a third centrifugation step (700g, 5 min, 4°C). Last, mitochondria were pelleted (8500g, 5 min, 4°C) and resuspended in 100 ml of macrophage inflammatory protein without BSA. Protein concentration of mitochondria was determined using Bradford reagent (Sigma-Aldrich) according to the manufacturer's instructions. Mitochondria were either immediately used (respirometry or in organello translation) or snap-frozen and stored at −80°C.

#### Blue native polyacrylamide gel electrophoresis Western blot analysis

Blue native polyacrylamide gel electrophoresis (BN-PAGE) was performed on the basis of the “NativePAGE Novex Bis-Tris Gel System” (Invitrogen) according to the manufacturer’s instructions. For analysis of mitochondrial supercomplexes, 10 µg of mitochondria was lysed with 4% of digitonin. After completion of lysis (15 min on ice), lysates were cleared (30 min, 20,000g, 4°C), and the resulting supernatant was loaded on a 4 to 16% bis-tris gradient gel. Subsequently, proteins were transferred to an Amersham Hybond polyvinylidene difluoride membrane (GE Healthcare) by Western blot and subsequently immunodecorated with indicated antibodies.

#### Cardiac tissue protein lysates

Homogenization of 25 mg of cardiac tissue samples in 400 ml of cold organ lysis buffer [50 M Hepes (pH 7.4), 50 mM NaCl, 1% Triton X-100 (v/v), 0.1 M NaF, 10 mM EDTA, 0.1% SDS (w/v), 10 mM Na-orthovanadate, 2 mM phenylmethylsulfonyl fluoride, 1 protease inhibitor cocktail Sigma-Aldrich), and 1  PhosSTOP phosphatase inhibitor cocktail (Roche)] was performed with the Precellys CK 14 (Bertin Technologies) (5000 rpm, 30 s). Cleared protein lysates (45 min, 20,000g, 4°C) were transferred into fresh tubes. Determination of protein concentration was performed with Bradford reagent (Sigma-Aldrich) according to the manufacturer’s instructions. Protein lysates were stored at 80°C.

#### SDS–polyacrylamide gel electrophoresis

Protein samples were dissolved in SDS-PAGE loading buffer [50 mM tris-HCl (pH 6.8), 2% SDS (w/v), 10% glycerol (v/v), 1% beta-mercaptoethanol, 12.5 mM EDTA, and 0.02% bromophenol blue] before denaturation. Depending on the required range of protein sizes, the proteins were separated on 8 to 15% acrylamide gels [stacking gel: 5% acrylamide-bisacrylamide (37.5:1), 12.5 mM tris-HCl, 0.1% SDS (w/v), 0.25% Ammonium persulfate (APS), and 0.25% Tetramethylethylenediamine (TEMED) (pH 6.8); separating gel: 8 to 15% acrylamide-bisacrylamide (37.5:1), 37.5 mM tris-HCl, 0.1% SDS (w/v), 0.1% APS, and 0.1% TEMED (pH 8.8)] in running buffer [25 mM tris-HCl, 250 mM glycine, and 0.1% SDS (w/v) (pH 8.3)].

**Immunoblot**

Transfer of proteins on a nitrocellulose membrane by Immunoblot was conducted in transfer buffer (30 mM tris-HCl, 240 mM glycine, 0.037% SDS, and 20% methanol) at 400 mA for 2 hours at 4°C. For a first evaluation of the transfer, shortly washed membranes (dH2O) were stained with Ponceau S solution (Sigma-Aldrich). Depending on the antibody requirements, destaining and blocking of membranes were performed for 1 hour either in 5% milk-PBST (Phosphate-Buffered Saline/Tween) or 3% BSA-TBST (Tris-Buffered Saline/Tween) on a gently shaking platform before subsequent immunodecoration with the indicated antibodies according to the manufacturer's instructions. Secondary horseradish peroxidase–coupled antibodies (1:2000) were incubated for 1 hour before detection by Pierce ECL Immunoblotting substrate (Thermo Fisher Scientific). Densitometry-based quantification of Western blots was performed with ImageJ and Image Studio Lite Software. For a complete list of antibodies see supplemental table 3.

**Total Collagen Assay**

Left ventricular collagen content was measured using the Abcam total collagen assay kit (ab222942, Abcam, Cambridge, United Kingdom) according to manufacturer's protocol. Briefly, 10 µg of freshly harvested left ventricles were homogenized in aqua dest. using a Peqlab Tissue Lyser (VWR, Radnor, PA, USA, #000-158) followed by an alkaline hydrolysis to yield free hydroxyproline, which was subsequently oxidized yielding chromophore that was detected using a spectrophotometer (Thermo Fisher Scientific, Waltham, MA, USA, Type 357, #357-902221T) at OD 560 nm.

**Proteomics**

A panel of 3072 plasma proteins was measured from the patients' heparin-plasma collected at initial and final visitation using the proximity extension assay method (Explore 3072, Olink, Uppsala, Sweden). Student's paired T-Test followed by Benjamini-Hochberg correction for multiple testing was applied. Visualization was performed using InstantClue freeware (https://github.com/hnolCol/instantclue, accessed October 25, 2022).

### Supplemental Figures Captions

Supplemental Figure 1: (a) Elisa for cardiac troponin I (CTnI) from plasma samples of indicated genotypes, (b) representative electron microscopy images depicting cardiac sarcomers of the indicated genotypes; scalebar = 1µm. (c) Total collagen content of left ventricles of the respective groups. Delta of left ventricular pressure (dP) in $Mlp^{-/-}$;$Mpo^{-/-}$ hearts in Langendorff perfusion at baseline and after treatment with (d) HSA and (e) MPO; verapamil and isoproterenol were used as controls for negative and positive inotropic stimulation. (f) Response in dP of Langendorff-perfused hearts relative to baseline for the indicated substances. For statistical analysis, One-way ANOVA followed by Tukey's post-hoc test was used. * p-value < 0.05, ** p-value < 0.01, *** p-value < 0.001, **** p-value < 0.0001.

Supplemental Figure 2: Elastase release from murine whole blood samples treated for 24h as indicated. Asterisks show significance against vehicle treatment. For statistical analysis, One-way ANOVA followed by Tukey's post-hoc test was used. **** p-value < 0.0001.

Supplemental Figure 3: (a) and (b) Immunoblots of the indicated proteins of mitochondrial and integrated stress response; HSC70 was used as a loading control (some loading control bands are displayed multiple times to facilitate visual comparison). (c) Digitonin-BN-PAGE showing indicated mitochondrial supercomplexes (SC) and complexes (Co). (d) Immunoblot of p-cMyBP-C and total cMyBP-C with HSC70 used as loading control.

Supplemental Figure 4: Quantification of immunoblots from supplemental figure 2a, b and e. For statistical analysis, One-way ANOVA followed by Tukey's post-hoc test was used. * p-value < 0.05, ** p-value < 0.01, *** p-value < 0.001.

Supplemental Figure 5: Extended analysis of echocardiography from MPO deficient $Mlp^{-/-}$ mice after two minutes of acute MPO infusion shows changes in LVEF (a), heart rate (b), end diastolic volume (c), cardiac output (d), stroke volume (e) and end systolic volume (f). For statistical analysis, One-way ANOVA followed by Tukey's post-hoc test was used. * p-value < 0.05, ** p-value < 0.01.

Supplemental Figure 6: Extended analysis of cardiac MRI and transthoracic echocardiography of HFrEF in patients diagnosed for DCM treated with an oral MPO-inhibitor (AZD4831) shows development from baseline (V1) to end of treatment (V8) for left ventricular enddiastolic volume (LVEDV, a), left ventricular endsystolic volume (LVESV, b), both LVEDV and LVESV indexed for body surface area (c, d), left ventricular enddiastolic diameter (LVEDD, c), left ventricular endsystolic diameter (LVESD, d), basal diameter of right ventricle (e) and tricuspid anular plane systolic excursion (f).

Supplemental Figure 7: Effect of 21 days treatment with Ang II in MPO deficient and MPO competent $Mlp^{-/-}$ mice on left ventricular parameters, i.e. LVEF (a), heart rate (b), enddiastolic volume (c), cardiac output (d), stroke volume (e), endsystolic volume (f). For statistical analysis, One-way ANOVA followed by Tukey's post-hoc test was used. * p-value < 0.05, *** p-value < 0.001.

Supplemental Figure 8: Volcano plot depicting a panel of 3072 plasma proteins measured in HFrEF patients prior to and after 12 weeks of treatment with MPO-inhibitor AZD4831 (a). Individual regulation of heart failure marker NTproBNP (b) IL-10 (c) and IL-13 (d). NPX = Normalized Protein eXpression, manufacturer's arbitrary unit which is in Log2 scale, i.e. a 1 NPX difference means a doubling of protein concentration.

| | **Patient 1** | **Patient 2** | **Patient 3** | **Patient 4** | **Patient 5** | **Mean** | **SD** |
|---|---|---|---|---|---|---|---|
| **Age** | 76-80 | 61-65 | 56-60 | 61-65 | 56-60 | 64,6 | 6,1 |
| **Sex** | male | male | male | female | male | | |
| **LVEF (%)** | 24 | 44 | 14 | 47 | 36 | 33 | 12,4 |
| **NTproBNP (U/l)** | 6842 | 1145 | 3009 | 361 | 348 | 2341 | 2450 |
| **6MWD (m)** | 184 | 460 | 460 | 276 | | 345 | 119,5 |
| **mPAP (mmHg)** | 36 | 28 | 25 | 25 | 26 | 28 | 4,1 |
| **Cause for DCM** | Idiopathic | Myocarditis | Idiopathic | Chemotherapy | Myocarditis | | |
| **Date of DCM diagnosis** | 2006 | 02/2021 | 01/2021 | 2013 | 2016 | | |

Supplemental Table 1: Baseline characteristics of patients treated with AZD4831

Abbreviations:

LVEF: Left ventricular ejection fraction

NTproBNP: N-terminal brain natriuretic peptide

6MWD: 6-minute walking distance

mPAP: mean pulmonary arterial pressure

DCM: Dilated cardiomyopathy

| **Patient 1** | | | | | | | | | |
|---|---|---|---|---|---|---|---|---|---|
| **Medication** | **Start Date** | **Screening** | **V1** | **V2** | **V3** | **V4** | **V5** | **V6** | **V8** |
| Bisoprolol 5 mg 1-0-1 | Oct-2010 | x | x | x | x | x | x | x | x |
| Torasemide 10 mg 1-1-0 | Oct-2010 | x | x | x | x | x | x | x | x |
| Spironolactone 25 mg 1-0-0 | Oct-2010 | x | x | x | x | x | x | x | o* |
| Ramipril 1,25 mg 1-0-1 | Oct-2010 | x | x | x | x | x | x | x | x |
| Apixaban 5 mg 1-0-1 | Mar-2015 | x | x | x | x | x | x | x | x |
| Dapagliflozine 5 mg 1-0-1 | Jan-2016 | x | x | x | x | x | x | x | x |
| Metformin 1000 mg 1-0-1 | Jan-2016 | x | x | x | x | x | x | x | x |
| Simvastatin 20 mg 0-0-1 | Aug-2020 | x | x | x | x | x | x | x | x |
| Amiodarone 200 mg 1-0-0 | Jan-2021 | x | x | x | x | x | x | x | x |
| Spironolactone 12,5 mg 1-0-0 | Jul-2021 | o | o | o | o | o | o | o | x |
| | | | | | | | | | *Reduced due to Hyperkalemia |
| **Patient 2** | | | | | | | | | |
| **Medication** | **Start Date** | **Screening** | **V1** | **V2** | **V3** | **V4** | **V5** | **V6** | **V8** |
| Bisoprolol 2,5 mg 1-0-1 | Feb-2021 | x | x | x | x | x | x | x | x |
| Sacubitril/Valsartan 24/26 mg 1-0-1 | Feb-2021 | x | x | x | x | x | x | x | x |
| Spironolactone 25 mg 1-0-0 | Feb-2021 | x | x | x | x | x | x | x | x |
| Phenprocumon n. INR | Feb-2021 | x | o | o | o | o | o | o | o |
| Dapagliflozine 10 mg 1-0-0 | Feb-2021 | x | x | x | x | x | x | x | x |
| Buproprion 150 mg 1-0-0 | Mar-2009 | x | x | x | x | x | x | x | x |

| | | | | | | | | | |
|---|---|---|---|---|---|---|---|---|---|
| Venlaflaxin 225 mg 1-0-0 | Feb-2009 | x | x | x | x | x | x | x | x |
| **Patient 3** | | | | | | | | | |
| **Medication** | **Start Date** | **Screening** | **V1** | **V2** | **V3** | **V4** | **V5** | **V6** | **V8** |
| Bisoprolol 5 mg 1-0-1 | Jan-1996 | x | x | x | x | x | x | x | x |
| Torasemide 50 mg 1-0-0 | Jan-2020 | x | x | x | x | x | x | x | x |
| Spironolactone 25 mg 1-0-0 | Jan-2020 | x | x | x | x | x | x | x | x |
| Sacubitril/Valsartan 24/26 mg 1-0-1 | Jan-2021 | x | x | x | x | x | x | x | x |
| Phenprocumon n. INR | Jan-1996 | x | x | x | x | x | x | x | x |
| Dapagliflozine 10 mg 1-0-0 | Mar-2021 | x | x | x | x | x | x | x | x |
| Metformin 1000 mg 1-0-1 | Mar-2010 | x | x | x | x | x | x | x | o |
| Simvastatin 20 mg 0-0-1 | Mar-2007 | x | x | x | x | x | x | x | x |
| Tamsulosin 0,4 mg 1-0-0 | Jan-2015 | x | x | x | x | x | x | x | x |
| Metformin 1000 mg 1-0-1/2 | Mar-2010 | o | o | o | o | o | o | o | x |
| **Patient 4** | | | | | | | | | |
| **Medication** | **Start Date** | **Screening** | **V1** | **V2** | **V3** | **V4** | **V5** | **V6** | **V8** |
| Metoprolol 23,75 mg 1-0-0 | Jan-2013 | x | x | x | x | x | x | x | x |
| Dapagliflozine 10 mg 1-0-0 | Jan-2021 | x | x | x | x | x | x | x | x |
| Spironolactone 25 mg 1-0-0 | Jan-2013 | x | x | x | x | x | x | x | x |
| Torasemide 10 mg 1-0-0 | Jan-2013 | x | x | x | x | x | x | x | x |
| Sacubitril/Valsartan 49/51 mg 1-0-1 | Jan-2019 | x | x | x | x | x | x | x | x |
| Ivabradine 5 mg 1-0-0 | Jan-2021 | x | x | x | x | x | x | x | x |
| Sitaglitpin 50 mg | Jan-2009 | x | x | x | x | x | x | x | x |

| | | | | | | | | | |
|---|---|---|---|---|---|---|---|---|---|
| 1-0-1 | | | | | | | | | |
| Metformin 1000 mg 1-0-1 | Jan-2009 | x | x | x | x | x | x | x | x |
| Omeprazole 20 mg 1-0-0 | Jan-2013 | x | x | x | x | x | x | x | x |
| L-Thyroxin 100 µg 1-0-0 | Jan-2005 | x | x | x | x | x | x | x | x |
| Salmeterol/Fluticason 50/500 µg | Jan-1995 | x | x | x | x | x | x | x | x |
| Atorvastatin 20 mg 0-0-1 | Jan-2013 | x | x | x | x | x | x | x | x |
| ASS 100 mg 1-0-0 | Jan-2013 | x | x | x | x | x | x | x | x |
| Paracetamol | Jul-2021 | o | o | o | o | o | o | x | x |
| Heparin s.c. | Jul-2021 | o | o | o | o | o | o | | |
| **Patient 5** | | | | | | | | | |
| **Medication** | **Start Date** | **Screening** | **V1** | **V2** | **V3** | **V4** | **V5** | **V6** | **V8** |
| Dapagliflozine 10 mg 1-0-0 | May-2021 | x | x | x | x | x | x | x | x |
| Torasemid 20 mg 1-0-0 | Jan-2016 | x | x | x | x | x | x | x | x |
| Bisoprolol 5 mg 1-0-1 | Jan-1996 | x | x | x | x | x | x | x | x |
| ASS 100 mg 1-0-0 | Jan-2021 | x | x | x | x | x | x | x | x |
| Clopidogrel 75 mg 1-0-0 | Jan-2021 | x | x | x | x | x | x | x | x |
| Sacubitril/Valsartan 24/26 mg 1-0-1 | Feb-2021 | x | x | x | x | x | x | o | o |
| Sacubitril/Valsartan 49/51 mg 1-0-1 | Feb-2021 | o | o | o | o | o | o | x | x |
| Atorvastatin 20 mg 0-0-1 | Jan-2021 | x | x | x | x | x | x | x | x |
| Pantoprazol 40 mg 1-0-0 | Jan-2021 | x | x | x | x | x | x | x | x |

Supplemental Table 2: Medication taken by patients treated with AZD4831 over the course of the study period.

X: taken

0: not taken

| **Name** | **Supplier** | **Cat. Nr.** |
|---|---|---|
| **AKT** | CST | 9272 |
| **p-AKT** | CST | 4060 |
| **ALDH18A1** | Sigma | HPA01260 |
| **ATP5A1** | Abcam | ab14748 |
| **BiP** | Santa Cruz | Sc-376768 |
| **CALNEXIN** | Calbiochem | 208880 |
| **CLPP** | Sigma | WH0008192M1 |
| **COX1** | Molecular Probes | 459600 |
| **COX4** | Molecular Probes | A21348 |
| **CPT1M** | alpha diagnostic | CPT1M11-A |
| **eIF2A** | Abcam | ab26197 |
| **p-eIF2a** | Abcam | ab32157 |
| **HSC70** | Santa Cruz | Sc-7298 |
| **Ire1a** | Santa Cruz | Sc-390960 |
| **LONP1** | Proteintech | 15440-1-AP |
| **MTHFD2** | Abcam | ab56772 |
| **mTOR** | CST | 2972 |
| **p-mTOR** | CST | 2971 |
| **MYBPC3** | Abcam | ab108522 |
| **p-MYBPC3(Ser-282)** | Enzo Life Sciences | ALX-215-057-R050 |
| **NDUFA9** | Molecular Probes | 459100 |
| **NDUFS1** | Proteintech | 12444-1-AP |
| **P62** | Abcam | ab91526 |
| **PYCR1** | Abcam | ab94780 |
| **SDHA** | Invitrogen | 459200 |
| **SHMT2** | Sigma Aldrich | AV46129 |
| **S6K** | CST | 2217 |
| **p-S6K** | CST | 2211 |
| **UQCRC1** | Molecular Probes | 459140 |
| **VDAC1** | CST | 4661 |

Supplemental Table 3: Complete list of antibodies used for immunoblots.

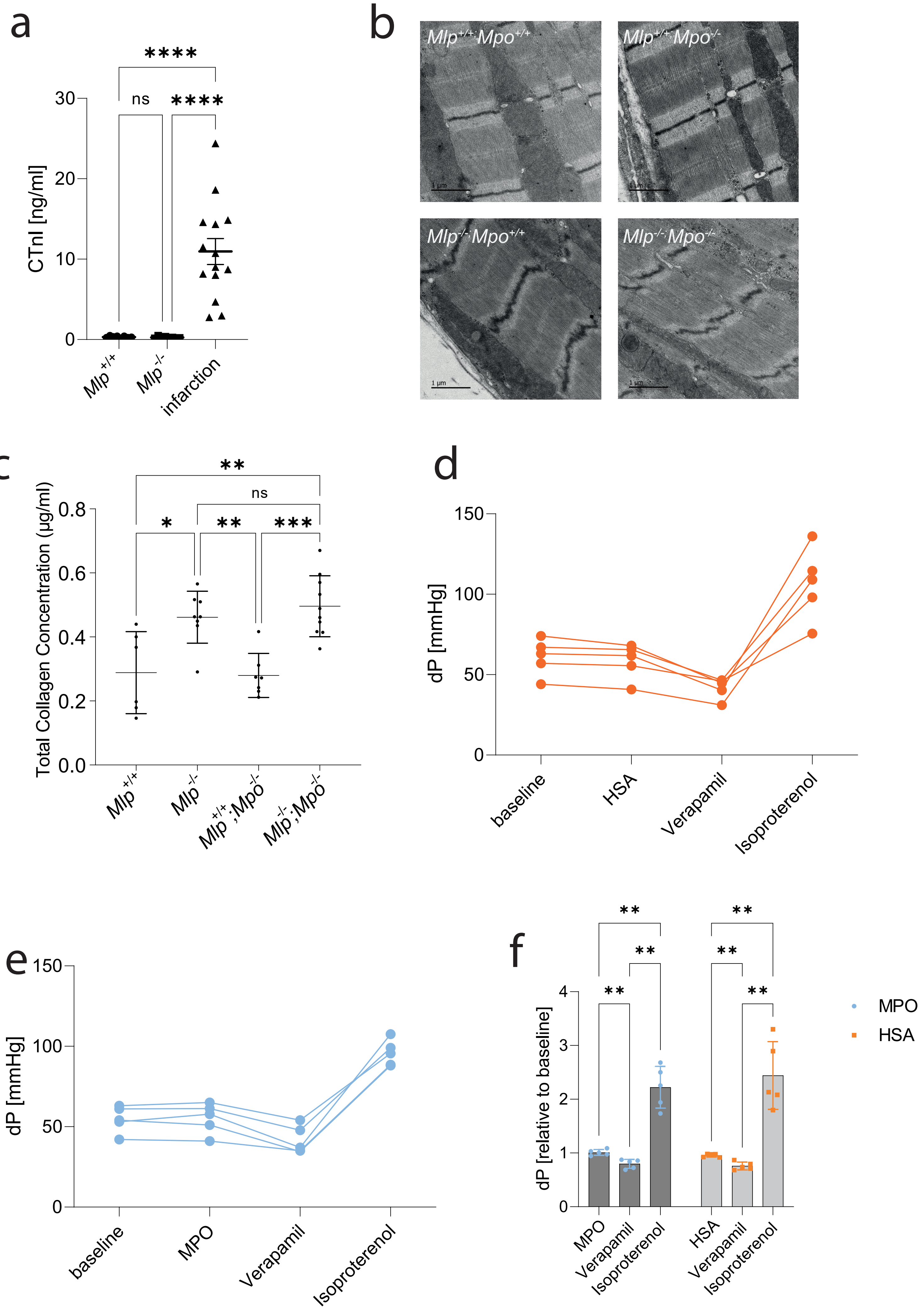

a
b
c
d
e
f
CTnI [ng/ml]
ns
****
Mlp+/+
Mlp-/-
infarction
Mlp+/+;Mpo+/+
Mlp+/+;Mpo-/-
Mlp-/-;Mpo+/+
Mlp-/-;Mpo-/-
1 µm
Total Collagen Concentration (µg/ml)
Mlp+/+;Mpo-/-
Mlp-/-;Mpo-/-
dP [mmHg]
baseline
HSA
Verapamil
Isoproterenol
MPO
dP [relative to baseline]

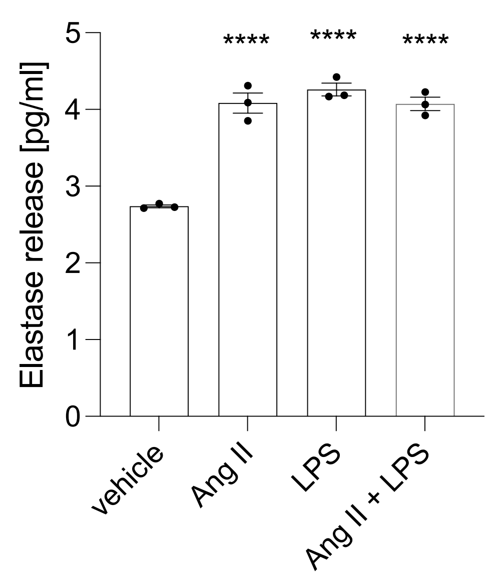

Elastase release [pg/ml]
0
1
2
3
4
5
****
****
****
vehicle
Ang II
LPS
Ang II + LPS

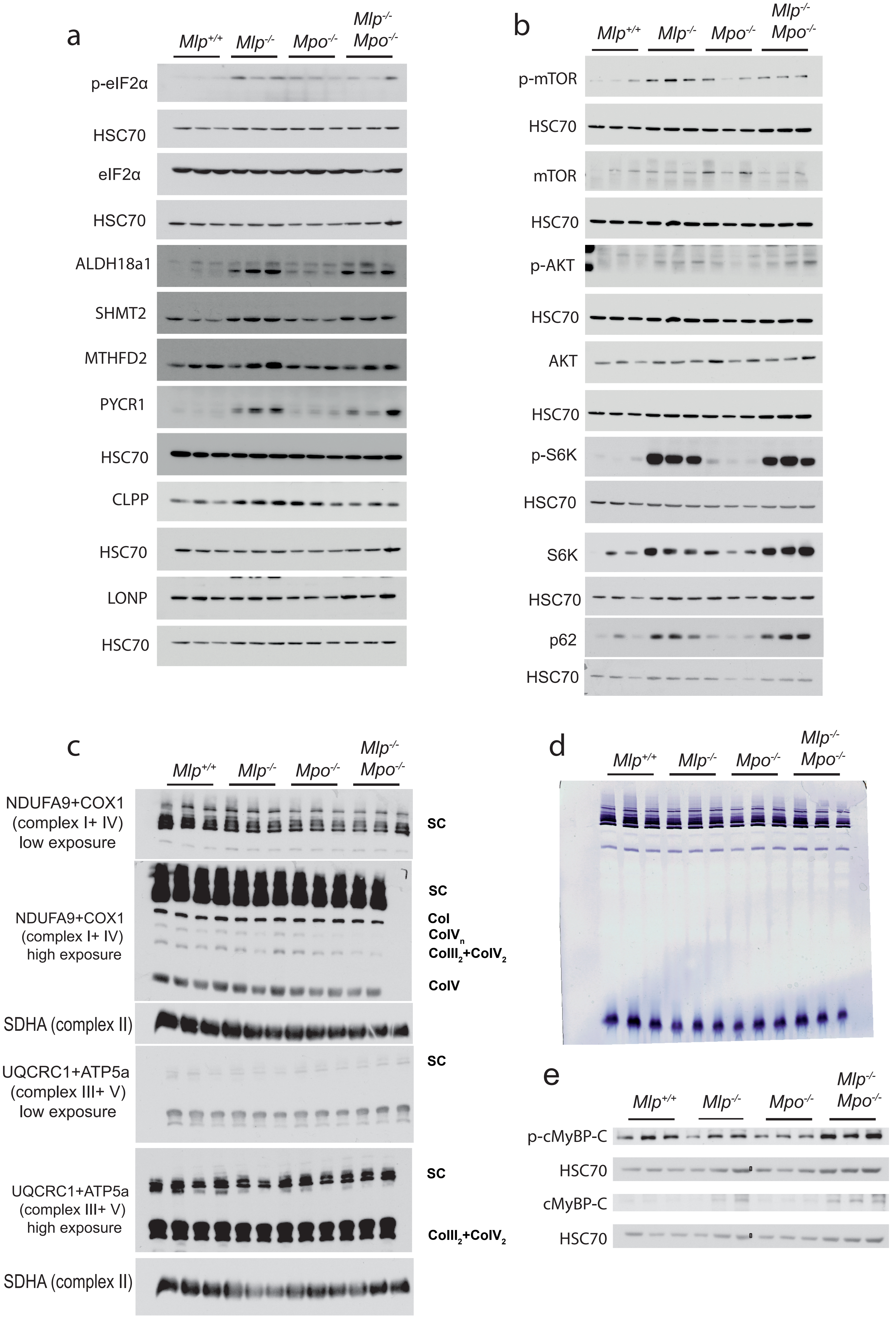
a
Mlp+/+
Mlp-/-
Mpo-/-
Mlp-/- Mpo-/-
p-eIF2α
HSC70
eIF2α
HSC70
ALDH18a1
SHMT2
MTHFD2
PYCR1
HSC70
CLPP
HSC70
LONP
HSC70
b
Mlp+/+
Mlp-/-
Mpo-/-
Mlp-/- Mpo-/-
p-mTOR
HSC70
mTOR
HSC70
p-AKT
HSC70
AKT
HSC70
p-S6K
HSC70
S6K
HSC70
p62
HSC70
c
Mlp+/+
Mlp-/-
Mpo-/-
Mlp-/- Mpo-/-
NDUFA9+COX1 (complex I+ IV) low exposure
SC
NDUFA9+COX1 (complex I+ IV) high exposure
SC
CoI
CoIVn
CoIII2+CoIV2
CoIV
SDHA (complex II)
UQCRC1+ATP5a (complex III+ V) low exposure
SC
UQCRC1+ATP5a (complex III+ V) high exposure
SC
CoIII2+CoIV2
SDHA (complex II)
d
Mlp+/+
Mlp-/-
Mpo-/-
Mlp-/- Mpo-/-
e
Mlp+/+
Mlp-/-
Mpo-/-
Mlp-/- Mpo-/-
p-cMyBP-C
HSC70
cMyBP-C
HSC70

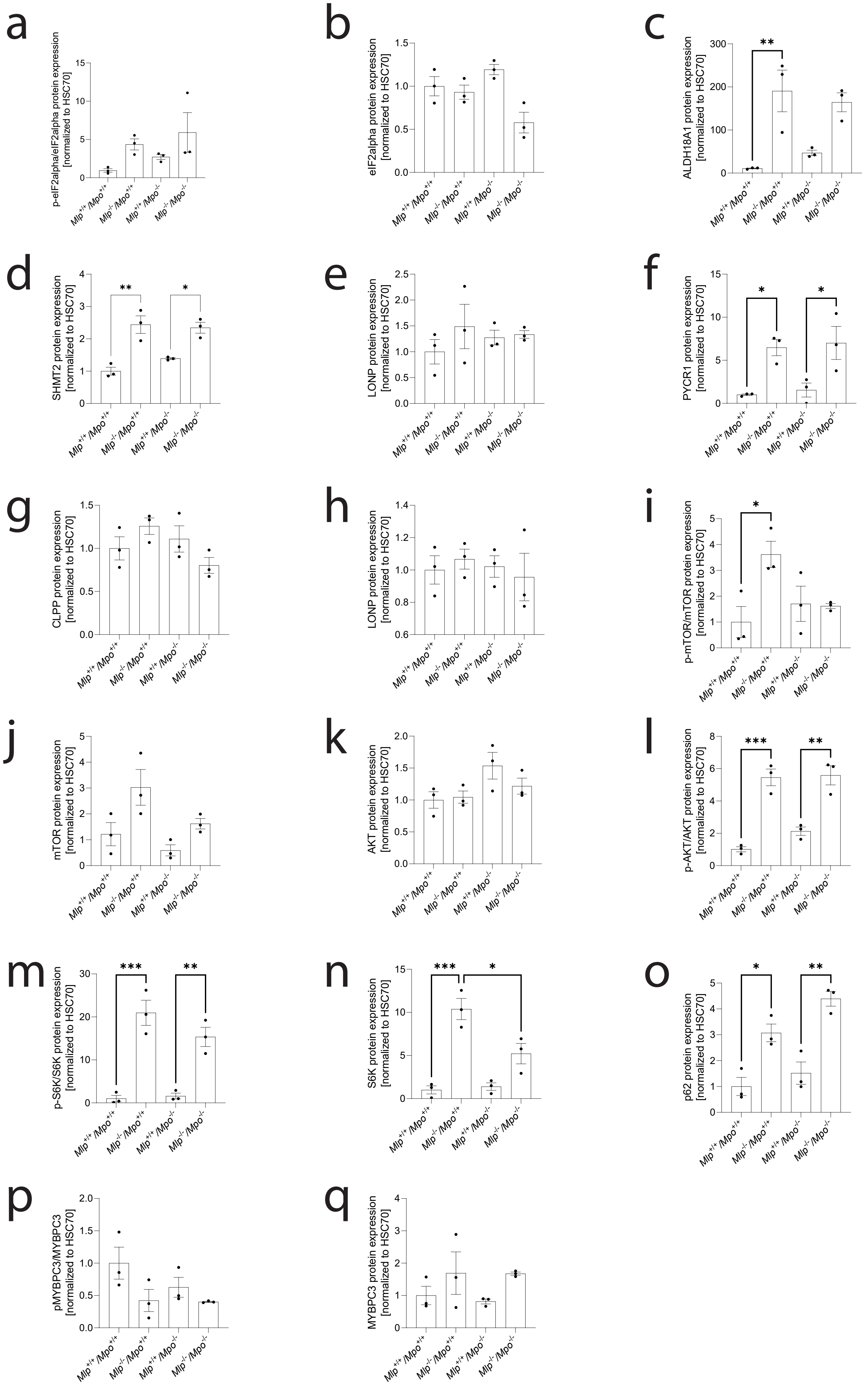
a
p-eIF2alpha/eIF2alpha protein expression [normalized to HSC70]
b
eIF2alpha protein expression [normalized to HSC70]
c
ALDH18A1 protein expression [normalized to HSC70]
d
SHMT2 protein expression [normalized to HSC70]
e
LONP protein expression [normalized to HSC70]
f
PYCR1 protein expression [normalized to HSC70]
g
CLPP protein expression [normalized to HSC70]
h
LONP protein expression [normalized to HSC70]
i
p-mTOR/mTOR protein expression [normalized to HSC70]
j
mTOR protein expression [normalized to HSC70]
k
AKT protein expression [normalized to HSC70]
l
p-AKT/AKT protein expression [normalized to HSC70]
m
p-S6K/S6K protein expression [normalized to HSC70]
n
S6K protein expression [normalized to HSC70]
o
p62 protein expression [normalized to HSC70]
p
pMYBPC3/MYBPC3 [normalized to HSC70]
q
MYBPC3 protein expression [normalized to HSC70]
Mlp+/+/Mpo+/+
Mlp-/-/Mpo+/+
Mlp+/+/Mpo-/-
Mlp-/-/Mpo-/-

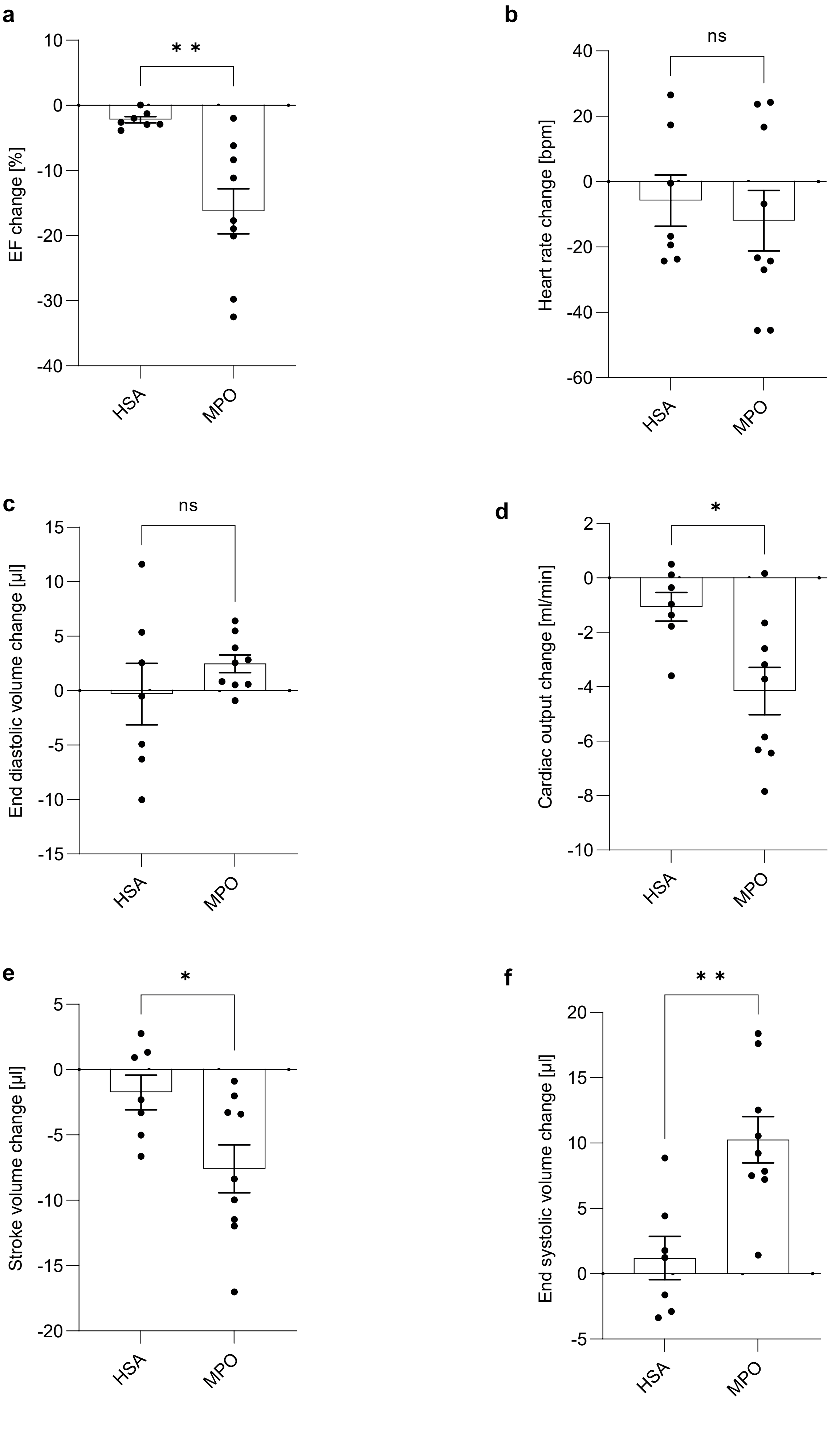
a
EF change [%]
10
0
-10
-20
-30
-40
**
HSA
MPO
b
Heart rate change [bpm]
40
20
0
-20
-40
-60
ns
HSA
MPO
c
End diastolic volume change [µl]
15
10
5
0
-5
-10
-15
ns
HSA
MPO
d
Cardiac output change [ml/min]
2
0
-2
-4
-6
-8
-10
*
HSA
MPO
e
Stroke volume change [µl]
5
0
-5
-10
-15
-20
*
HSA
MPO
f
End systolic volume change [µl]
20
15
10
5
0
-5
**
HSA
MPO

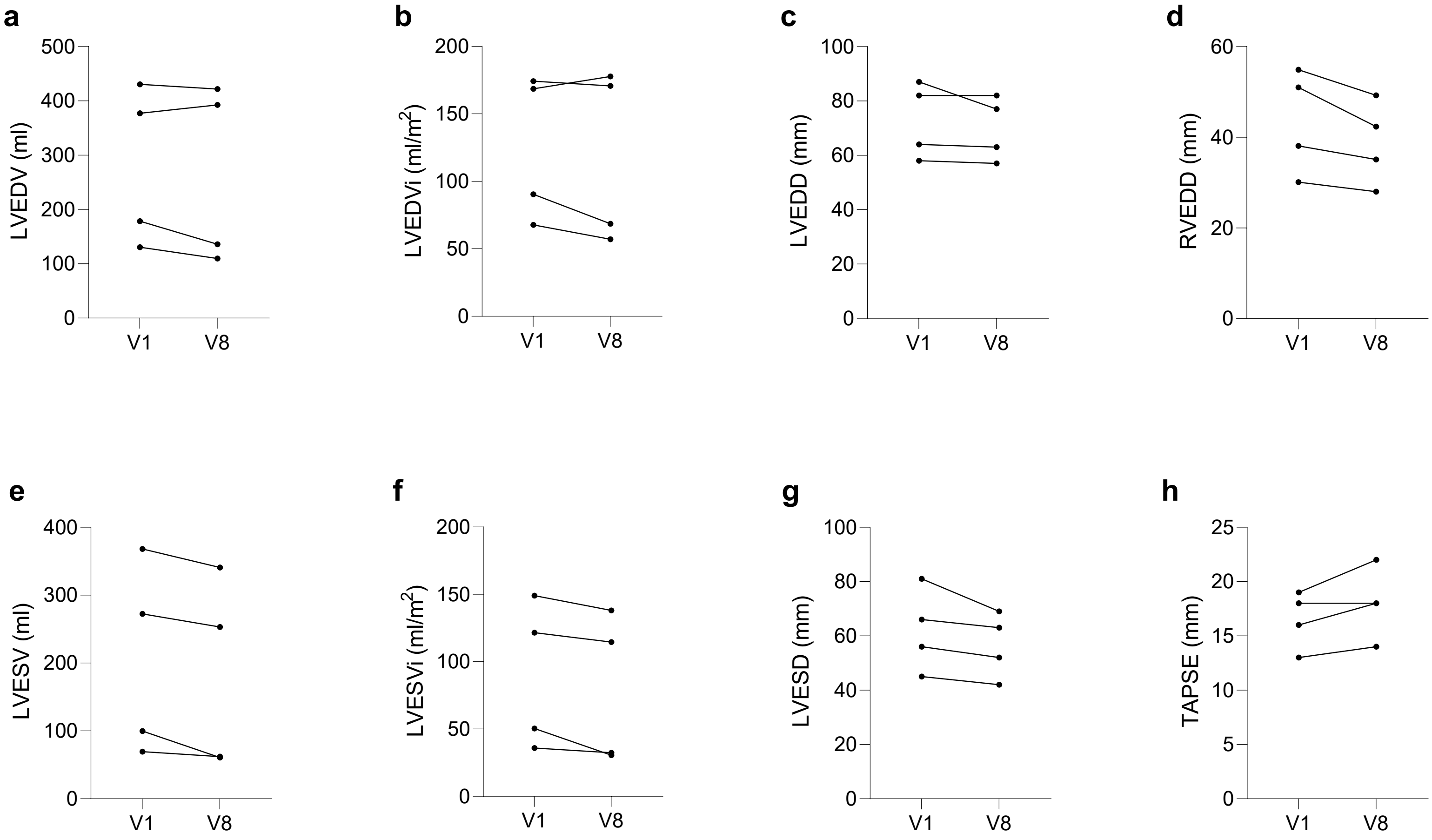

a
LVEDV (ml)
500
400
300
200
100
0
V1
V8
b
LVEDVi (ml/m2)
200
150
100
50
0
V1
V8
c
LVEDD (mm)
100
80
60
40
20
0
V1
V8
d
RVEDD (mm)
60
40
20
0
V1
V8
e
LVESV (ml)
400
300
200
100
0
V1
V8
f
LVESVi (ml/m2)
200
150
100
50
0
V1
V8
g
LVESD (mm)
100
80
60
40
20
0
V1
V8
h
TAPSE (mm)
25
20
15
10
5
0
V1
V8

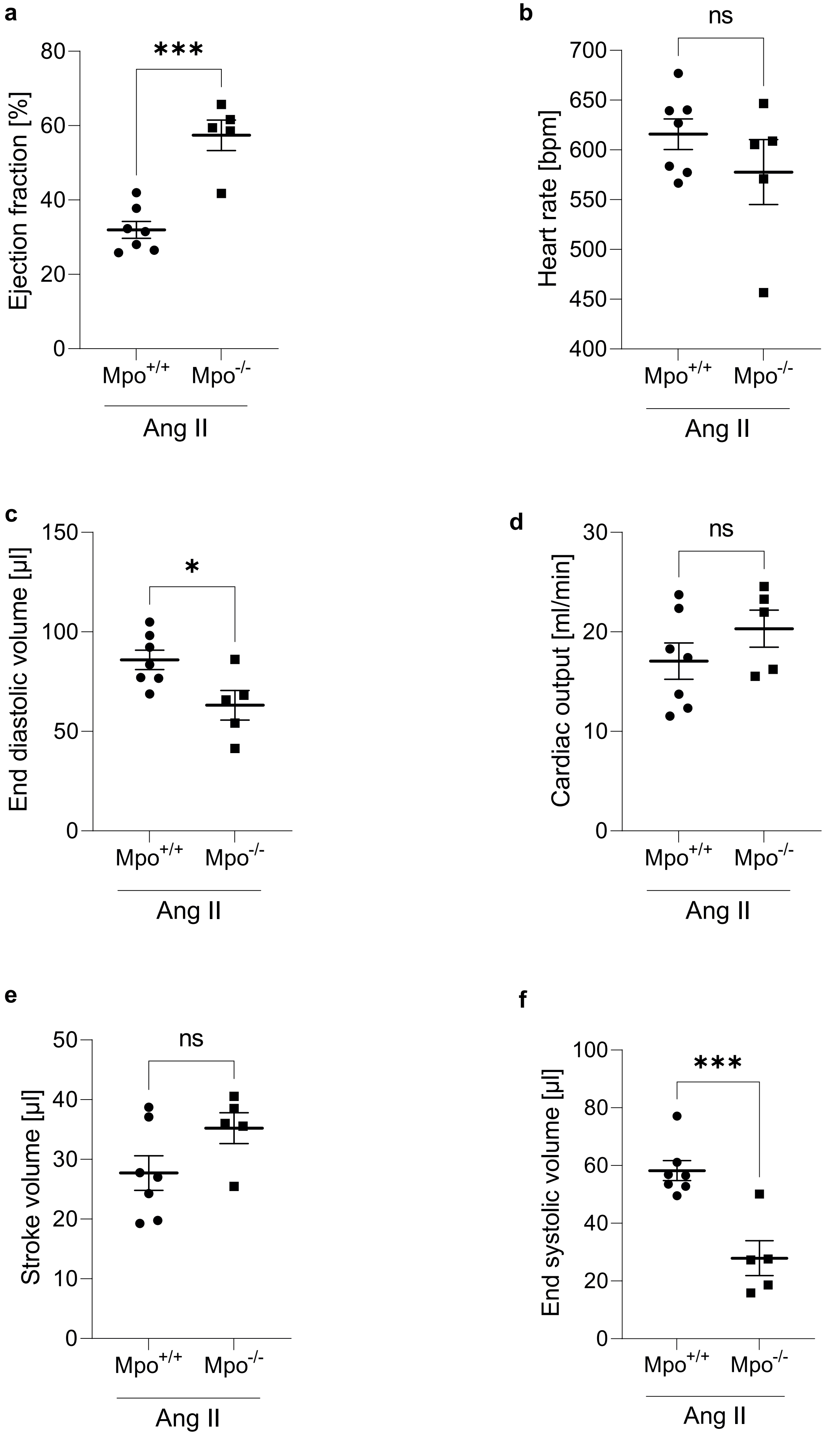
a
***
Ejection fraction [%]
80
60
40
20
0
Mpo+/+
Mpo-/-
Ang II
b
ns
Heart rate [bpm]
700
650
600
550
500
450
400
Mpo+/+
Mpo-/-
Ang II
c
*
End diastolic volume [µl]
150
100
50
0
Mpo+/+
Mpo-/-
Ang II
d
ns
Cardiac output [ml/min]
30
20
10
0
Mpo+/+
Mpo-/-
Ang II
e
ns
Stroke volume [µl]
50
40
30
20
10
0
Mpo+/+
Mpo-/-
Ang II
f
***
End systolic volume [µl]
100
80
60
40
20
0
Mpo+/+
Mpo-/-
Ang II

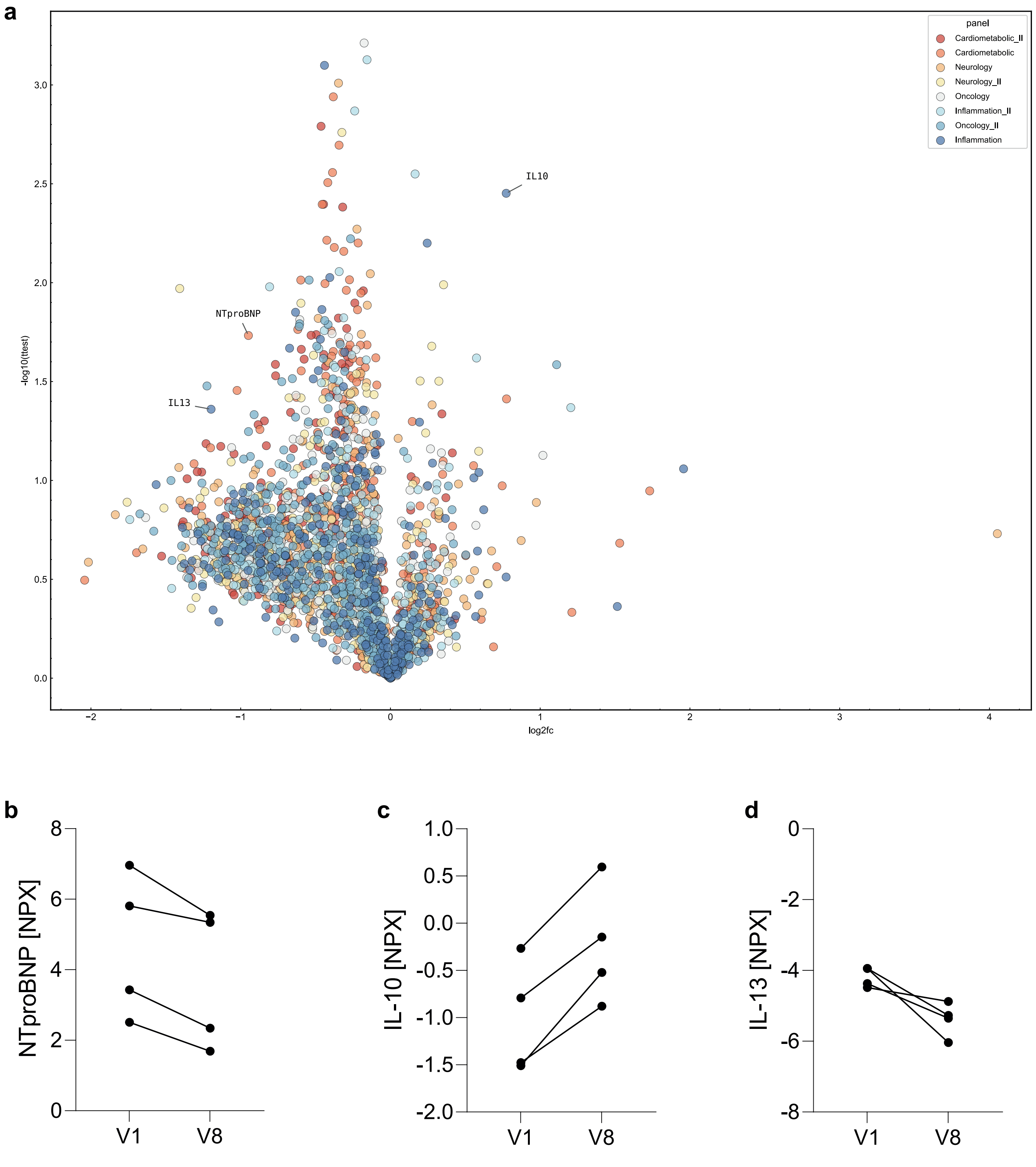
a
panel
Cardiometabolic_II
Cardiometabolic
Neurology
Neurology_II
Oncology
Inflammation_II
Oncology_II
Inflammation
IL10
NTproBNP
IL13
-log10(ttest)
log2fc
b
NTproBNP [NPX]
V1
V8
c
IL-10 [NPX]
V1
V8
d
IL-13 [NPX]
V1
V8
